# Development of a new Aerosol Barrier Mask for mitigation of spread of SARS-CoV-2 and other infectious pathogens

**DOI:** 10.1101/2021.02.11.21251593

**Authors:** Karam Abi Karam, Piyush Hota, S. Jimena Mora, Amelia Lowell, Kelly McKay, Xiaojun Xian, Bhavesh Patel, Erica Forzani

## Abstract

The COVID-19 pandemic has caused huge impact on public health and significantly changed our lifestyle. This is due to the fast airborne oro-nasal transmission of SARS-CoV-2 from the infected individuals. The generation of liquid aerosolized particles occurs when the COVID-19 patients speak, sing, cough, sneeze, or simply breathe. We have developed a novel aerosol barrier mask (ABM) to mitigate the spread of SARS-CoV-2 and other infectious pathogens. This Aerosol Barrier Mask is designed for preventing SARS-CoV-2 transmission while transporting patients within hospital facilities. This mask can constrain aerosol and droplet particles and trap them in a biofilter, while the patient is normally breathing and administrated with medical oxygen. The system can be characterized as an oxygen delivery and mitigation mask which has no unfiltered exhaled air dispersion. The mask helps to prevent the spread of SARS-CoV-2, and potentially other infectious respiratory pathogens and protects everyone in general, especially healthcare professionals.

## 1. Introduction

As of February 5^th^, 2021, the Word Health Organization reported more than 104 million confirmed cases of COVID-19, including more than 2.2 million deaths globally.^1^ These substantial numbers are the consequence of the transmission of the Severe Acute Respiratory Syndrome Coronavirus 2 (SARS-CoV-2). It is transmitted by droplets and aerosols generated by infected individuals during their exhalation.^2^ An individual infected with COVID-19, would potentially infect others by natural human actions such as breathing, speaking, coughing, sneezing, etc.^3,4,5^ Healthcare professionals are the most vulnerable population since they are caring for infected patients.^6,7^ Considering the severity of the pandemic, it is important to reduce and control the spread of the virus.^8^ Our group has developed an Aerosol Barrier Mask (ABM) that significantly reduces the airborne transmission of virus-containing respiratory droplets from in-patients during their transportation in hospital facilities. The design of the mask also includes an oxygen fitting allowing its administration to the patient. ABM is considered an innovative and efficient system designed particularly to protect healthcare professionals. This Aerosol Barrier Mask is proved to be very effective in mitigating the spread of SARS-CoV-2 in hospital settings.

## 2. Materials and Methods

The three subjects involved in the test study participated voluntarily and signed informed consents.

The tests were carried out by Arizona State University (ASU) researchers at Mayo Simulation Centers at Phoenix, Arizona in April 2021. The study was approved by the Institutional Review Board of Arizona State University (IRB reference protocols # STUDY00006547).

### 2.1 Mask Design and Components

The mask was redesigned and retrofit using an existing single-use mask from the Breezing Pro™ metabolic tracker.^9^ The mask is equipped with two connectors as observed in Fig. 1A and B. The central coupler is an outlet port that connects to the main flow Anti-Bacterial/Viral Filter, which further connects to an appropriately designed fan to suction out the aerosols generated from the wearer (flow rate: 3.6-4.3 CFM at zero static pressure). The second connector is an oxygen port, sealed with a medical-grade adhesive, allowing the administration of medical oxygen. The mask has a comfortable and adjustable strap to hold it in position on the patient’s head. The fan is powered by a 5V portable and a rechargeable battery pack, and it can be reused for multiple times.

**Figure 1.**
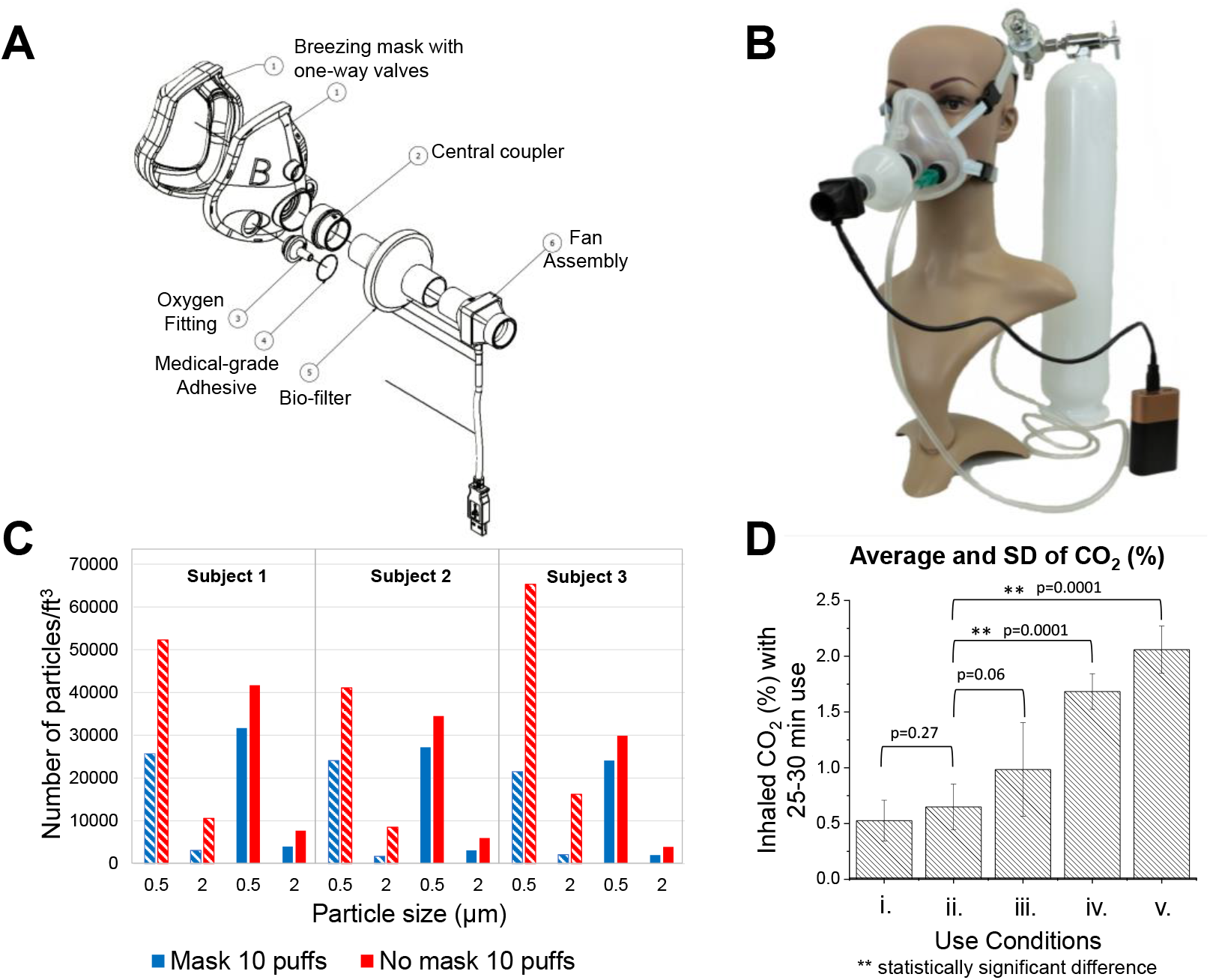
**A, B)** Mask designed for a safer transportation of COVID-19 patients in healthcare facilities. The ABM has an outlet port with a bio-filter connected to a fan for negative pressure and a port for oxygenation. **C)** Plots of maximum number of aerosol particles/feet^3^ *vs* particle size measured by simulated coughing and sneezing at different distances from a subject. (hatched columns correspond to 1.3 ft and solid to 2 ft) with and without the ABM (blue and red, respectively). The average of the maximum number of particles/feet^3^ for the 0.5 µm and 2 µm particle size corresponding to the baseline is represented by the dashed and dash-dotted line respectively. **D)** The graph shows the inhaled CO_2_ levels of the ABM system under different conditions − i. O_2_ with Fan OFF, ii. O_2_ with Fan ON, iii. ambient air via an emergency inhalation valve with fan ON, iv. ambient air via an emergency inhalation valve with fan OFF, v. CO_2_ concentration with a reference N95 mask in ambient air.

### 2.2 Particles Count

To quantify the number of aerosols, a commercial laser particle counter, Dylos DC1100 (Dylos Corporation, CA) was used. This air quality monitor detects particles between 0.5 and 2.5 microns. The monitors were installed at three distinct locations away from the subject − 1.3 ft towards the left, 2 ft towards the right, and 13 ft longitudinally away (data not shown). The aerosols generated by coughing and sneezing simulations were accomplished by ten puffs with a sprayer at the subject’s mouth level. The resulting concentration of aerosol particulates were studied with and without the ABM system.

### 2.3 Comparison of Inhaled Carbon Dioxide Measurement

An infrared CO_2_ VacuMed analyzer, silver edition Model 17630, was used to measure the concentration of the corresponding gas accumulated by breath using the ABM. The assessments of CO_2_ levels were conducted in 5 different conditions:

i. ABM with filter, 15 LPM of oxygen, and fan turned OFF
ii. ABM with filter, 15 LPM of oxygen, and fan turned ON
iii. ABM with filter, fan turned ON, with no oxygen tank
iv. ABM with filter, fan turned OFF, with no oxygen tank
v. An N-95 mask, with ambient air for comparison

It is important to note, with the absence of oxygen supply, ambient air was available to the user through the one-way inhalation valve on the mask. The data was collected using a Bluetooth Data Acquisition Module -BTH-1208LS manufactured by Measurement Computing. Measurements were taken every 5 minutes with a 1-minute interval for a total time of 30 minutes. The CO_2_ inhalation average and standard deviation were calculated using the acquired data set.

## 3. Results

The mask is specially designed to provide safe movement/transportation of COVID-19 patients who generate contaminated aerosols via coughing, sneezing, or by use of their vocal cords. In addition, the system can be used when the patient is inside an imaging system such as a CT scanner without contaminating the surroundings. The ABM is a light and comfortable single-patient use mask (≤100 g) that is easy to place on the person’s face.

Figure 1C shows the plot of the maximum number of aerosol particles per cubic feet measured during the experiments (typically at a time lapse of 20 minutes) for particle sizes corresponding to 0.5 µm and 2 µm at different distances from 3 subjects with and without the ABM system. The assembly of mask, filter, and fan enabled to mitigate almost 100% of the dispersed aerosol particles effectively. On the other side, the generation of aerosols was detected specifically for short distances. It is important to note that the ventilation of the room has a big influence on aerosol detection in short distances. For example, in this case-study, more aerosols were detected at 2 feet distance rather than 1.3 feet.

Fig. 1D shows the inhaled carbon-dioxide levels using the ABM for different conditions as discussed in *section 2.3*. It is evident that the fully functional aerosol mitigation mask shows better breathing conditions than an N95 mask supplied with ambient air. Even at other conditions, i.e. if one of the components of the system fails (fan or oxygen), the results assured relatively safer conditions for the wearer. Importantly, the characteristics of the fan are relevant and must sustain a flow compatible with the subject’s breathing rate (e.g., breathing peak flow, frequency, end-tidal volume, and average exhalation rate) in addition to the oxygen therapy’s flow which is dependent on the type of oxygen delivery.

## 4. Conclusions

An Aerosol Barrier Mask has been developed to prevent the spread of SARS-CoV-2 while transporting infected in-patients through hallways, elevators, and other places within hospital facilities. It has been shown that the ABM can mitigate close to a 100% of the aerosols and droplets generated by the wearer, which helps protect the individuals in proximal contact with the infected patients. Moreover, there is no carbon-dioxide accumulation inside the mask, outside the normal breathing range when the patient is administrated with oxygen. The ABM is a new alternative to help the mitigation of COVID-19.

## Data Availability

All data is in our possession and available if is required

## Acknowledgments

The authors acknowledge SEMTE, ASU, Mayo Clinic, Biodesign Institute, and NIH R03 funding from NIBIB (EB027336-02)

